# Impact of Praziquantel Distribution on the Epidemiology of Paragonimiasis Among School-Aged Children: A Cross Sectional Study in the Most Endemic Focus in Cameroon

**DOI:** 10.1101/2024.11.27.24318100

**Authors:** Fabrice Zobel Lekeumo Cheuyem, Jean-Brice Fomeni Toubue, Henri Donald Mutarambirwa, Guy Roger Pilo Ndibo

## Abstract

**Background:** Paragonimiasis is a parasitic disease affecting humans and other mammals, caused by infestation with lung flukes of the genus *Paragonimus*. The peri-urban area of Kumba is co-endemic for paragonimiasis and schistosomiasis. The National Schistosomiasis and Soil-Transmitted Helminthiasis Control Program distributes praziquantel annually for schistosomiasis control. The primary objective of this study was to describe the shellfish cooking habits among schoolchildren and to assess the impact of praziquantel mass distribution on the paragonimiasis burden in the most endemic focus in Cameroon.

**Methods:** We carried out a cross-sectional descriptive study from November 2013 to March 2014. Pupils of five government primary schools in five villages around Kumba underwent both clinical and parasitological investigations in search of signs and symptoms of paragonimiasis. The Chi square and Fisher exact tests were used to compared proportions. *p*-values<0.05 were considered statistically significant.

**Results:** We recruited 175 children, comprising 96 females and 79 males, with a sex ratio (F/M) of 1.2. Their ages ranged from 5 to 15 years, with a mean age of 10 ± 3.74 years. The age group 6-10 years was the most represented. All participants reported consuming crabs. The prevalence of paragonimiasis among the school children was 0.57% (95% CI: 0.0143%-3.143%). This result was statistically significantly lower than that of the last study conducted ten years prior, which used the same diagnostic method in this area (12.3%). Slightly more than half of the students (58.9%) reported preparing their crabs by boiling, and two-thirds (69%) estimated the cooking time to be more than 30 minutes. More than two-thirds (78%) of the children who estimated the cooking time to be more than 30 minutes consumed shrimp with their family. Pulmonary symptoms were the most common, with cough being the most prevalent. Notably, we did not register any cases of hemoptysis. Coinfections were also found in the population studied.

**Conclusions:** The prevalence of paragonimiasis has decreased in this focus, known to be the most endemic for this disease in Cameroon. This is due to the large distribution of praziquantel in this area and probably to changes from unhealthy cooking habits to healthier ones.

## Background

Paragonimiasis, also known as endemic hemoptysis, is one of the most important foodborne anthropozoonoses caused by one or more trematodes of *Paragonimus spp*. The disease is endemic in several countries in Asia, Africa and South America [5]. In 2005, an estimated 23.2 million people worldwide were infected with paragonimiasis, including 5 million severe infection that resulted in and death [6].

The definitive hosts of this parasite comprise humans and a diverse range of wild animals, primarily belonging to the *Canidae* (canine) and *Felidae* (feline) families. [7]. The life cycle of this parasite involves a complex series of intermediate hosts, including numerous species of snails (Gastropoda) that serve as the first intermediate host, and various crustaceans, such as crabs (Brachyura) and crayfish (Astacoidea), which act as the second intermediate host [8].

Most often, humans become infected by eating raw or undercooked freshwater crustaceans containing metacercariae, the infesting larval stage of the parasite [9]. More rarely, humans can become infected by ingesting raw or undercooked pork or wild boar, which serve as paratenic hosts [10]. Elsewhere, notably in Korea, people enjoy traditional dishes made from raw freshwater crabs and crayfish, which could lead to significant *Paragonimus spp*. infestation [11,12]. Nevertheless, some patients diagnosed with paragonimiasis reveal that they have never had to eat freshwater crabs or crayfish. It is therefore possible that they have eaten food contaminated by the hands or cooking utensils of people who have recently handled these crustaceans [13]. In line with guidelines from the Food and Drug Administration, shellfish should be cooked to an internal temperature of at least 145°F (63°C) for at least 30 minutes to ensure food safety [14].

The acute phase of infestation (parasite invasion and migration) is clinically manifested by diarrhea, abdominal pain, fever, cough, urticaria, hepatosplenomegaly and hypereosinophilia [15]. On the other hand, cough, hemoptysis, chest pain and chest X-ray abnormalities reflect the chronic phase of the disease [5]. These abnormalities may persist for several years, even after adequate treatment [16].

In Cameroon, there are several known foci of paragonimiasis. Such foci include the Kupé Montain [17–19], the Mundani in the South-West Region [20], the Mbam and Nyong in the Centre Region, the Ntem in the South Region [18]. The previous study to assess the morbidity burden of paragonimiasis was conducted in Kumba with a prevalence of 12.3% [21].

Since 2009, as part of the National Schistosomiasis and Soil-Transmitted Helminthiasis Control Program, the Kumba Health District, where prevalence of schistosomiasis was estimated to be 10%, has benefited from mass distribution campaigns of praziquantel to all children aged 5 to 15 [22–24]. This mass distribution could have had an impact on the distribution of paragonimiasis, which is also sensitive to praziquantel [25]. We conducted this study with the aim of describe the shellfish cooking habit among schoolchildren and the impact of praziquantel mass distribution of paragonimiasis burden in the most endemic focus in Cameroon.

## Methods

### Study Design and Period

To achieve our objectives, we conducted a cross-sectional observational study from February to March 2014.

### Study Site and Population

The study population comprised pupils from government primary schools in five villages surrounding Kumba town, specifically Barombi-Kang, Ediki, Mabonji, Etam, and Teke. The present study was carried out in the departments of Kupé-Muanenguba and Mèmè, representing two of the six departments of the Southwest Region. The department are crossed by the Mungo and Mèmè rivers. Both departments are located in the forest zone of the Southwest Region, about 100 km from the Atlantic Ocean, with geographic coordinates 44° 41’ 32” N, 91° 12’ 35” E. The climate is tropical, with two seasons. Kumba is the largest city in the Southwest Region. The primary economic activity of the study population is agriculture, encompassing food and cash crop farming, as well as plantain agriculture operated by the Cameroon Development Corporation [26].

### Study Participants and Sampling

The study included all pupils who presented pulmonary symptoms of the disease (cough, hemoptysis, chest pain) whether or not associated with any other sign or symptom of paragonimiasis, who had lived in the area for at least one year, and whose parents or guardians had given their consent. Assent was also obtained from the study participants prior to sample collection with headmasters serving as witness.

### Sample Size and Sampling Technique

The minimum sample size was computed using the following formula: 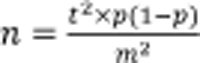 [27]. Where n: minimum sample size required to obtain significant results for a given event, for a given level of risk; t: the value of the reduced centered normal distribution for a 95% confidence level which is 1.96; p: probability of occurrence (12.3%) [26]; m: margin of error (usually set at 5%). 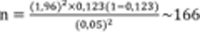. We therefore needed 166 participants for the results to be statistically valid. A cluster random sampling technique was used to select the classes from which participants were eventually enrolled.

### Clinical Investigation

A questionnaire was used to collect clinical data. These questions included sociodemographic characteristics, cooking habits, medical examination to investigate symptoms of paragonimiasis, including cough, hemoptysis, and chest pain.

### Parasitological analysis

Participants were then provided with containers to collect sputum and stool samples, which were subsequently mixed with sodium azide and transported to the base laboratory in Yaounde for analysis. Sputum samples were treated with caustic soda, centrifuged, and examined under a light microscope for the presence of *Paragonimus spp*. eggs. Stool samples were analyzed using the Ritchie concentration technique to detect *Paragonimus spp.* eggs and other co-infecting parasites. Patient diagnosed with paragonimiasis were treated with praziquantel at a dose of 3 × 25 mg/kg body weight per day for three consecutive days [25].

### Data Analysis

Statistical results were generated using Epi Info version 3.5.1, Microsoft Office Excel version 2010. Confidence intervals (CI) were estimated at the 95% level. Independent groups were compared using chi-squared (*X*^2^) or Fisher’s exact test for proportions. The difference between independent proportions was considered statistically significant if their respective confidence intervals did not overlap. A *p*-value<0.05 was considered statistically significant.

## Results

## 1. Sociodemographic Characteristic of Participants

A total of 175 participants were selected from five village government primary schools in Kumba Health District, namely Barombi-Kang, Ediki, Etam, Mabonji and Teke. There was no significant difference between the number of participants enrolled from different schools (*p*-value=0.70) (Table 1).

**Table 1.** Distribution of study participants by village.

| School | Count (n) | Frequency (%) | p-value<sup>1</sup> |
| --- | --- | --- | --- |
|  |  |  | 0.70 |
| Barombi-Kang | 30 | 17 |  |
| Mabonji | 37 | 21 |  |
| Teke | 40 | 23 |  |
| Ediki | 35 | 20 |  |
| Etam | 33 | 19 |  |
| Total | 175 | 100 |  |

<sup>1</sup>Chi-square test

The sex ratio was (F/M) was 1.2. Participants were aged 05-15 years, with a mean of 10 years (mean = 10.19 ± 3.49 years for males and 9.84 ± 3.92 years for females). The 6-10 age group was the most represented (57%). Most of the pupils recruited were from the North-West Region (58%) (Table 2).

**Table 2.** Sociodemographic characteristics of study participants (*n*=175)

| Characteristic | Count (n) | Frequency (%) |
| --- | --- | --- |
| **Age (years)** |  |  |
| ≤5 | 4 | 2 |
| 6-10 | 100 | 57 |
| 11-14 | 69 | 40 |
| ≥15 | 2 | 1 |
| **Gender** |  |  |
| Female | 96 | 55 |
| Male | 79 | 45 |
| **Region** |  |  |
| Centre | 1 | 1 |
| Littoral | 1 | 1 |
| North | 2 | 1 |
| North-West | 102 | 58 |
| South-West | 65 | 37 |
| West | 4 | 2 |

## 2. Paragonimiasis Determinants

### 2.1. Shellfish Consumption Frequency

All school children who participated in this study ate crabs. Most of them reported a weekly consumption 128 (73%). There was no significant difference between age group (*p*=0.376) (Table 3).

**Table 3.** Frequency of crab consumption by age group among pupils.

| Age group | Frequency *n*(%) |  |  | Total (100%) | *p*-value<sup>1</sup> |
| --- | --- | --- | --- | --- | --- |
|  | Weekly | Monthly | Occasionally |  |  |
| ≤5 | 3(75) | 0(0) | 1(25) | 04 | 0.376 |
| 6–10 | 74(74) | 4(4) | 22(22) | 100 |  |
| 11–14 | 49(75) | 9(75) | 11(75) | 69 |  |
| ≥15 | 2(100) | 0(0) | 0(0) | 02 |  |
| Total | 128(73) | 13(7) | 34(20) | 175 |  |

<sup>1</sup>Fisher exact probability test

### 2.2. Shellfish Cooking Habit

Most respondents reported that they usually boiled (n=103; 59%) or fried (n=65; 37%) the crabs. A small proportion reported roasting before eating (n=7; 4%). Among those who estimated the cooking time of shellfish to be more than 30 minutes, more than two-thirds (78%) reported consuming it with their family, while 16% reported eating it with friends. However, the difference between these two groups was not statistically significant (*p*=0.389) (Table 4).

**Table 4.**
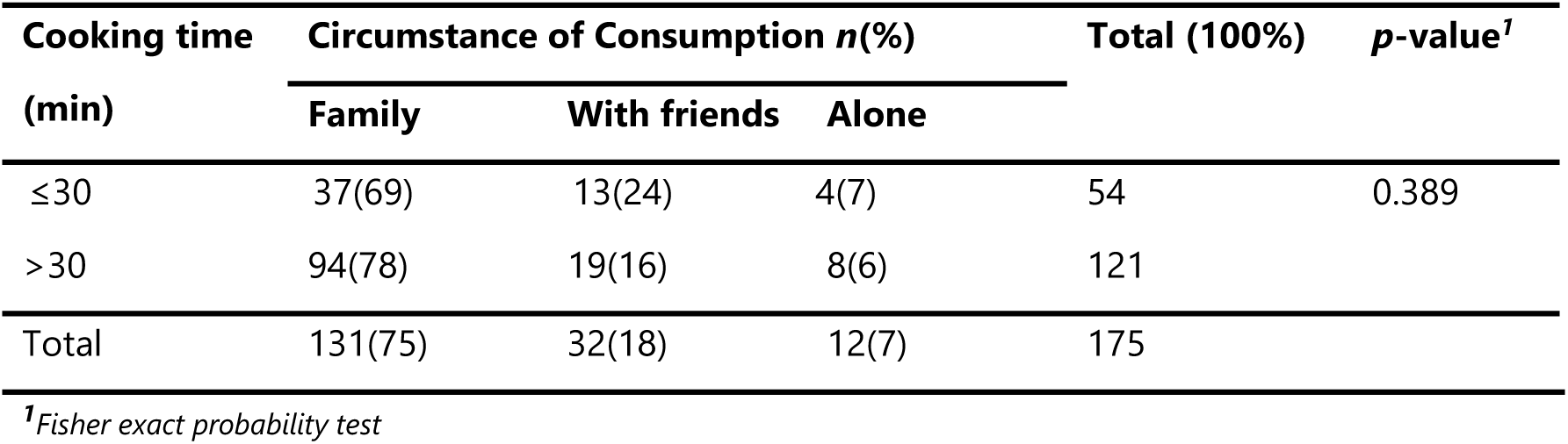
Crab cooking and consumption habits among school children.

This epidemiological survey also showed that when they were at school, where they spent almost 70% of their day, 82% of the participants reported disposing of their excreta in the toilets provided for this purpose, whereas only 18% reported doing so in the surrounding area.

### 2.3. Mass Campaign Participation

Most of the respondents (80%) reported having received a dose of praziquantel distributed during the last mass campaign of schistosomiasis prevention within school settings.

## 3. Clinical Profile

All participants presented with a cough. Chest pain (30.9%) and other non-specific signs of paragonimiasis, particularly abdominal pain (55.43%), were also common among the schoolchildren. Notably, there were no cases of hemoptysis (Figure 2).

**Fig. 1.**
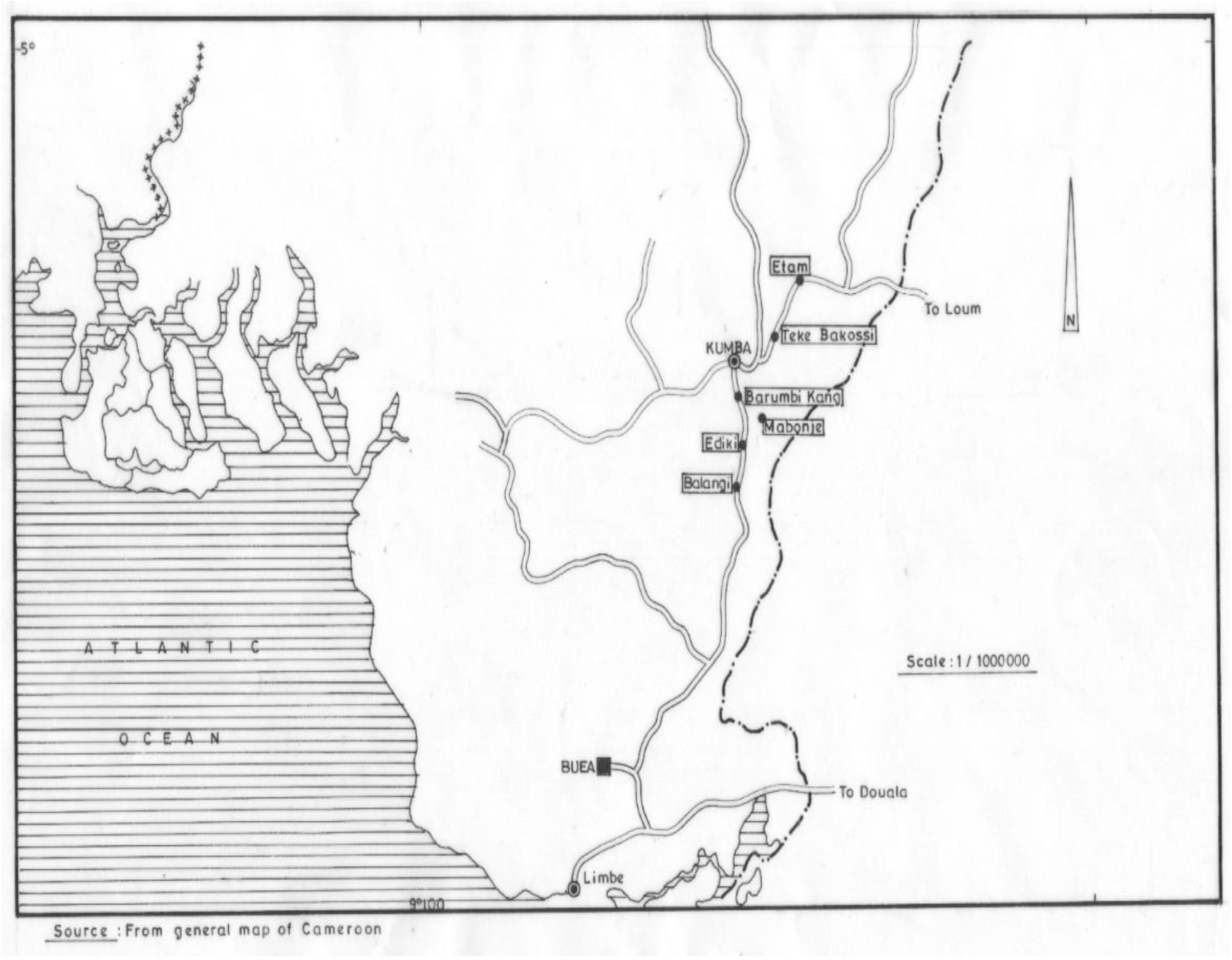
Map of the study site

**Fig. 2.**
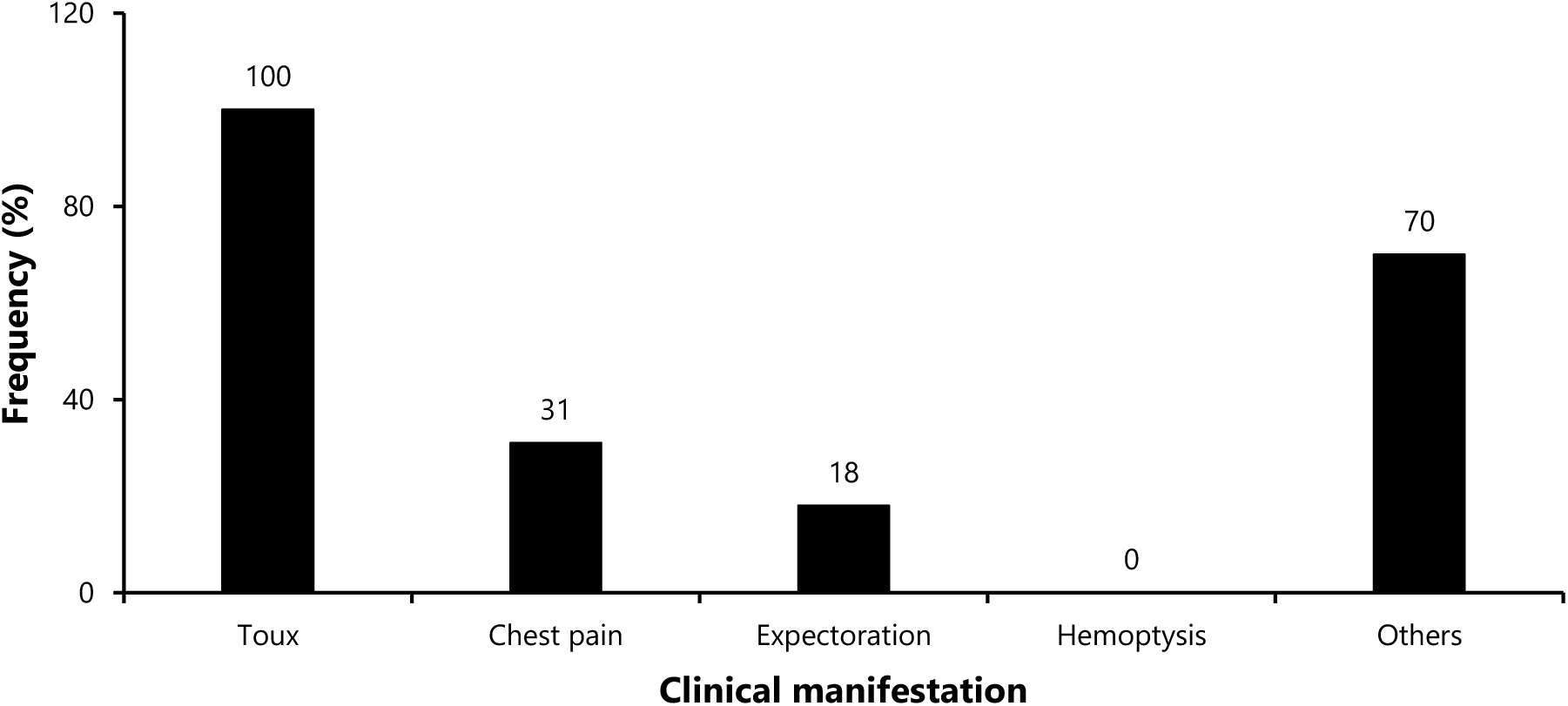
Clinical profile of participants (*n*=175)

## 4. Sputum and Stool Examination Findings

One case of the disease was found among all recruited school children (prevalence=0.57%; 95%CI: 0.0143%; 3.143%). These *Paragonimus spp*. eggs were detected in the sputum of an Etam schoolboy, aged 09 from the North-West Region. On examination, he presented with cough, chest pain and rusty sputum.

No *Paragonimus spp.* eggs were found in any of the stool samples. Nevertheless, eggs of soil-transmitted parasites were identified among 40 pupils (23%). Such parasites included mainly *Ascaris lumbricoides* (56%), followed by *Trichuris trichiura* (35%). Boys were not significantly more infested than girls by soil-transmitted parasites (*p*=0.458). (Table 5).

**Table 5.**
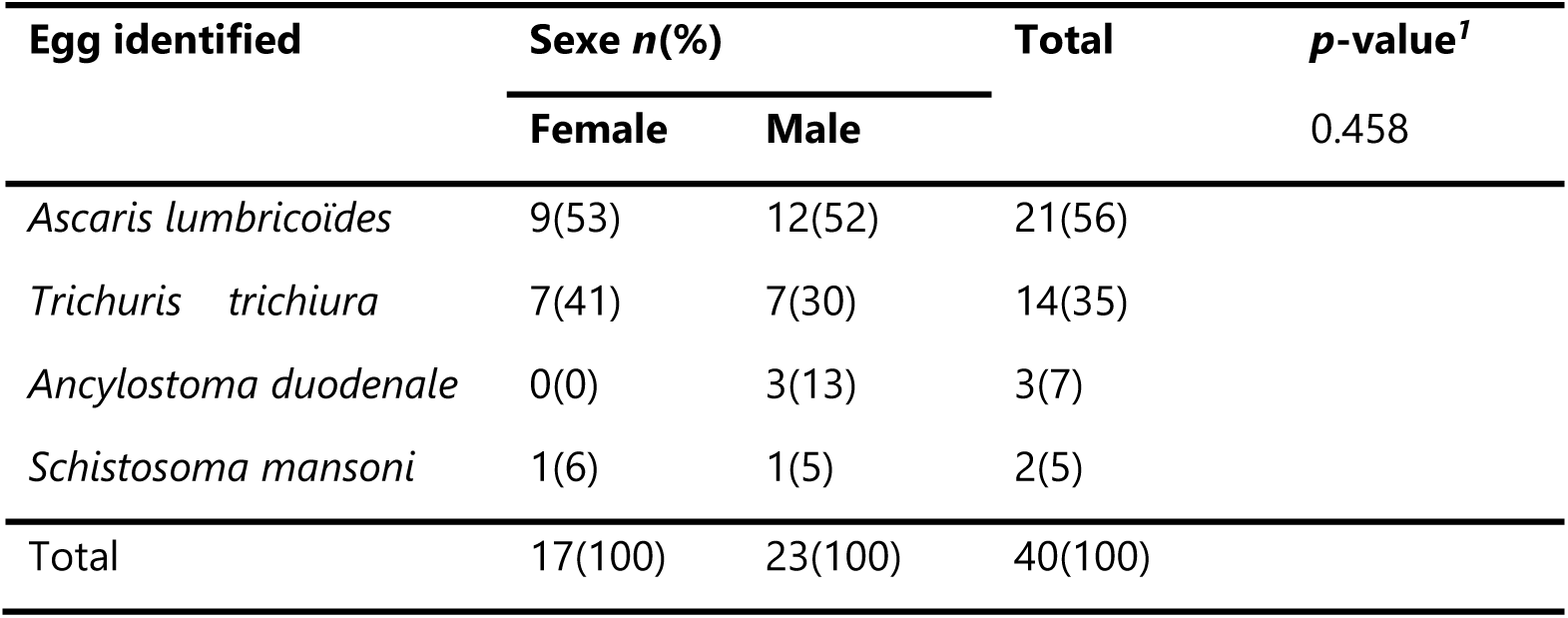
Profile of parasites identified in the pupils’ stools.

## Discussion

We were able to select primary schools similar to those visited in a previous study by Moyou *et al.* in 2003 and obtain an almost similar age distribution of participants, in order to obtain comparable results [21]. Analysis of stool samples for the diagnosis of paragonimiasis remains useful in children, as they usually swallow the product of bronchial secretions. However, the sensitivity of sputum examination remains superior to that of stool [28]. Moreover, Moyou *et al.* in 2003 found more eggs in sputum than in stool [21]. This could explain why the only positive case was found in sputum.

Studies have shown that a cure rate of 71-75% was achieved after the first day of praziquantel treatment [29]. According to the Kumba Health District, mass distribution campaigns of praziquantel (Biltricide® 600 mg tablets) in single doses ranging from 600 to 2400 mg, depending on height, have been carried out in the five years prior to this study. Our survey showed that 82% of children aged 05-15 years benefited from this treatment between May and June 2013. We believe that this wide annual distribution of praziquantel has contributed to a significant decrease in the prevalence of paragonimiasis from 12.3% [8.8%; 16.5%] in 2003 to a prevalence of 0.57% [0.01%; 3.1%] in 2014 [21]. Our result corroborate findings in India [30]. Furthermore, we found only two cases (1.14%) of *S. mansoni* schistosomiasis in our sample. However, it should be noted that treatment is only provided once a year, and participants can be reinfected during the year by eating undercooked crabs containing *Paragonimus spp*. metacercariae. Therefore, we believe that changes in people’s culinary habits may also have played a role in the decline of this disease.

All participants in this study consumed crabs, with 73% consuming them weekly. These data show that crab consumption habits in the peri-urban villages of Kumba have not changed since the study by Moyou *et al.* in 2003 [21]. The constancy of this high shellfish consumption can be explained by the fact that crabs remain widely available in the rivers surrounding the various villages. Moreover, these crustaceans remain culturally accepted delicacies, especially in Regions known to be endemic for paragonimiasis as illustrated in a study conducted in Nigeria [31].

Most of the participants reported consuming boiled crabs (58.9%) and 69% estimated their crab cooking time at over 30 minutes. Whereas, according to the study conducted by Moyou *et al.* in 2003, only 18.42% of schoolchildren reported boiling their crabs, and all associated the change in shell color as a sign of cooking; the required cooking time was obviously not reached [21]. It should also be noted that, to eliminate parasites such as *Paragonimus spp.* metacercariae, crabs must be cooked in water at over 63°C for at least 30 minutes [25]. We can therefore affirm that the evidence of compliance with these hygiene rules in the preparation of crabs by a significant proportion of the participants in our study could also explain the statistically significant decrease in the prevalence of paragonimiasis in this endemic area, despite the high consumption of crabs noted in both studies. In fact, the nonobservance of hygienic rules in the preparation of these crustaceans is responsible for the persistence or even the re-emergence of paragonimiasis in areas previously declared eradicated [32].

If we consider that family consumption of crabs implies better supervision of children by older children to ensure sufficient cooking time, and possibly cleaning of utensils after use, we believe that this may have played a role in the decline in the prevalence of paragonimiasis in this Region since the 2003 study [21]. In fact, 78% of the participants who estimated the cooking time of crabs to be more than 30 minutes usually consumed crabs with their family.

In addition, several studies have been carried out on paragonimiasis in this area by researchers [4].This would probably have raised awareness of the disease in the communities, particularly among the village chiefs usually contacted during community research, as well as primary schools’ headmasters, of the risks of eating undercooked crabs, and of the need to adopt healthier culinary habits. A similar hypothesis was underlines by Ollivier *et al.* to explain the drop in the prevalence of pulmonary paragonimiasis in southern Cameroon [33]. In Nigeria, the study conducted in a context of re-emerging paragonimiasis in certain Regions of the country, showed that 73% of participants did not clean utensils after preparing crabs [32]. Elsewhere, however, particularly in South Korea, the prevalence of paragonimiasis fell substantially following the impact of certain factors, namely the adoption of healthy eating habits and the cessation of the use of crabs in juices used in traditional medicine [34,35].

Our study revealed that 80% of people deposited their faeces in latrines. In fact, four out of five schools visited during our study had well-equipped, clean latrines, while only one had none. This high proportion of participants using latrines to deposit their feces would probably have reduced the dissemination of *Paragonimus spp.* eggs into the environment from subjects carrying the parasite, and consequently reduced the infestation of crabs by metacercariae. This would have contributed to lowering the prevalence of paragonimiasis in this area. On the other hand, in Nigeria, a study found that 87% of individuals deposited their feces in nearby rivers and bushes. These unsanitary habits are thought to be responsible for the re-emergence of paragonimiasis in this country [32]. It is important to note that the transmission of paragonimiasis is maintained by pollution of the water on which snails and crustacean vectors feed through feces and sputum.

Cough (100%) and chest pain (31%) were the most frequent presumptive symptoms of paragonimiasis. The low specific prevalence of paragonimiasis (0.57%) may be explained by the fact that its symptoms can also be found in other viral, bacterial, mycotic or even parasitic pathologies, notably during larval migrations in Löfflerian syndromes.

We also noted several cases of co-infections with other intestinal parasites, including *Ascaris lumbricoides, Trichuris trichiura, Ancylostoma duodenale,* and *Schistosoma mansoni*. These co-infections could explain the high frequency of abdominal pain among other symptoms of the disease in the participants. However, the prevalences of these parasites remain lower than those reported by Moyou *et al.* in 2003 [21]. This decrease may be attributed to the massive distribution of deworming products (Albendazole) during various national deworming campaigns conducted in the Health District in recent years.

The absence of cases of hemoptysis among the participants is further evidence of a marked decline in paragonimiasis. Indeed, studies which have detected considerable prevalences of paragonimiasis over the last few decades in this Region found cases of hemoptysis in 11-42% of participants [4,21,36].

## Limitations

The measurement bias in this study lies in the detection of eggs in samples, which is not possible in the pre-patent phase of infection or in cases of extrapulmonary paragonimiasis [37]. Nevertheless, this study remains comparable to the one carried out in 2003 [26], as we used the same diagnostic methods on the same type of population. Stool and sputum samples were examined at the IMPM laboratory in Yaoundé, as the technical platform did not allow us to do so at the Kumba District Hospital laboratory, nor was there a qualified analyst capable of reanalyzing and confirming the results obtained.

We encountered difficulties, especially among children under five years of age, who were sometimes unable to answer the questions correctly, or were unable to provide stool and/or sputum samples. This limited their participation in the study. A study reassessing the parasitic load of *Paragonimus spp.* metacercariae in crabs collected from the rivers of the villages in our study would further support these results. Nevertheless, it would have been interesting for these participants to have undergone serological tests of high diagnostic value such as ELISA, whose sensitivity and specificity can reach 100% [38], in order to support the alternative hypothesis with greater precision.

## Conclusions

The prevalence of schistosomiasis in this known focus decreased significantly from 2003 to 2014. Mass distribution campaigns of praziquantel have had an impact on the observed decrease in prevalence. Despite the high consumption of crabs, changes in hygiene measures in crab preparation may also have played a role in the decrease in paragonimiasis prevalence. We therefore suggest that mass distribution of antihelminthic drugs be continued. Further investigation should be conducted in the departments of Mèmè and Kupé-Muanenguba to re-evaluate the serological prevalence of paragonimiasis, including determining the prevalence of crabs infected with *Paragonimus spp.* metacercariae. The population should be sensitized to the fight against paragonimiasis and other intestinal parasites through the adoption of healthier hygiene measures.

## Abbreviations

CI: Confidence Interval

## Declarations

### Author contributions

Study design & conception: FZLC; Data collection: FZLC; Data analysis, visualization and interpretation: FZLC; Drafting of original manuscript: FZLC; Critical revision of the manuscript: FZLC, J-BFT, HDM and GRPN; Final approval of the manuscript: All authors.

### Ethical Approval Statement

The study was approved by the Ethics Committee of the Faculty of Medicine of Yaounde. Administrative approval was obtained from the Regional Delegate of Public Health of the Southwest Region, the Subdivisional Delegate of Basic Education of the Meme Department, the District Medical Officer of Kumba, and the directors of the various educational institutions selected for the study. Informed consent was obtained from the parents or guardians of the schoolchildren participating in the study. The confidentiality, anonymity and autonomy of the research participants were respected throughout the study. All procedures were carried out in accordance with the relevant guidelines of the Declaration of Helsinki.

### Consent for publication

Not applicable.

### Availability of data and materials

All data generated or analyzed during this study are included in this published article.

## Data Availability

All data produced in the present work are contained in the manuscript

## Acknowledgments

We are grateful to the headmasters, and the school children of the visited villages for their cooperation. This study received support from the Cameroon Institute of Medical Research and Study of Medicinal Plants (IMPM).

## References

1. Control of foodborne trematode infections. Report of a WHO Study Group. World Health Organ Tech Rep Ser. 1995; 849:1–157.

2. Keiser J, Utzinger J. Emerging foodborne trematodiasis. Emerg Infect Dis. 2005;11(10):1507–14.

3. Voelker J, Vogel H. 2 new Paragonimus species from West Africa: Paragonimus africanus and Paragonimus uterobilateralis (Troglotrematidae; Trematoda). Z Für Tropenmedizin Parasitol. 1965;16(2):125–48.

4. Nkouawa A, Okamoto M, Mabou AK, Edinga E, Yamasaki H, Sako Y, et al. Paragonimiasis in Cameroon: molecular identification, serodiagnosis and clinical manifestations. Trans R Soc Trop Med Hyg. 2009;103(3):255–61.

5. Jiang Y xin, Li G qiang, Pan C jing, He Z qiu, Wang C, Mu Q ru, et al. Pediatric paragonimiasis: a retrospective analysis of cases from a county in south-west China. Front Pediatr. 2023;11.

6. Fürst T, Keiser J, Utzinger J. Global burden of human food-borne trematodiasis: a systematic review and meta-analysis. Lancet Infect Dis. 2012;12(3):210–21.

7. Esteban-Sánchez L, García-Rodríguez JJ, García-García J, Martínez-Nevado E, de la Riva-Fraga MA, Ponce-Gordo F. Wild Animals in Captivity: An Analysis of Parasite Biodiversity and Transmission among Animals at Two Zoological Institutions with Different Typologies. Animals. 2024;14(5):813.

8. Doanh PN, Tu LA, Van Hien H, Van Duc N, Horii Y, Blair D, et al. First intermediate hosts of Paragonimus spp. in Vietnam and identification of intramolluscan stages of different Paragonimus species. Parasit Vectors. 2018;11(1):328.

9. Ortiz JB, Uys M, Seguino A, Thomas LF. Foodborne Helminthiasis. Curr Clin Microbiol Rep. 2024;11(3):153–65.

10. Chai JY. Chapter 23 - Paragonimiasis. In: Garcia HH, Tanowitz HB, Del Brutto OH, editors. Handbook of Clinical Neurology. Elsevier. 2013. p. 283–96.

11. Song JH, Dai F, Bai X, Kim TI, Yang HJ, Kim TS, et al. Recent Incidence of Paragonimus westermani Metacercariae in Freshwater Crayfish, Cambaroides similis, from Two Enzootic Sites in Jeollanam-do, Korea. Korean J Parasitol. 2017;55(3):347–50.

12. Cho AR, Lee HR, Lee KS, Lee SE, Lee SY. A Case of Pulmonary Paragonimiasis with Involvement of the Abdominal Muscle in a 9-Year-Old Girl. Korean J Parasitol. 2011;49(4):409–12.

13. Nakamura-Uchiyama F, Mukae H, Nawa Y. Paragonimiasis: a Japanese perspective. Clin Chest Med. 2002;23(2):409–20.

14. Centers for Disease Control and Prevention (CDC). Human paragonimiasis after eating raw or undercooked crayfish --- Missouri, July 2006-September 2010. MMWR Morb Mortal Wkly Rep. 2010;59(48):1573–6.

15. Zarrin-Khameh N, Citron DR, Stager CE, Laucirica R. Pulmonary Paragonimiasis Diagnosed by Fine-Needle Aspiration Biopsy. J Clin Microbiol. 2008;46(6):2137–40.

16. Procop GW. North American Paragonimiasis (Caused by Paragonimus kellicotti) in the Context of Global Paragonimiasis. Clin Microbiol Rev. 2009;22(3):415–46.

17. Zahra. Paragonimiasis in the Southern Cameroons: a preliminary report. West Afr Med J. 1952; 25:75–82.

18. Ripert C, Carrie J, Ambroise-Thomas P, Baecher R, Kum NP, Same-Ekobo A. Epidemiologic and clinical study of paragonimosis in Cameroon. Results of niclofolan treatment. Bull Société Pathol Exot Ses Fil. 1981;74(3):319–31.

19. Kum PN, Nchinda TC. Pulmonary paragonimiasis in Cameroon. Trans R Soc Trop Med Hyg. 1982;76(6):768–72.

20. Moyou-Somo R, Enyuong P, Kouanmouo J, Dinga JS, Couprie B, Ripert C. Etude de la paragonimose dans 5 villages du département de la Mèmè (Sud-Ouest Cameroun). Résultats du traitement par le praziquantel. Rev Sc Tech Yaoundé. 1983;6–7:125–331.

21. Moyou-Somo R, Kefie-Arrey C, Dreyfuss G, Dumas M. An epidemiological study of pleuropulmonary paragonimiasis among pupils in the peri-urban zone of Kumba town, Meme Division, Cameroon. BMC Public Health. 2003;3(1):40.

22. Campbell SJ, Stothard JR, O’Halloran F, Sankey D, Durant T, Ombede DE, et al. Urogenital schistosomiasis and soil-transmitted helminthiasis (STH) in Cameroon: An epidemiological update at Barombi Mbo and Barombi Kotto crater lakes assessing prospects for intensified control interventions. Infect Dis Poverty. 2017;6(1):49.

23. Tchuem Tchuenté LA, Kamwa Ngassam RI, Sumo L, Ngassam P, Dongmo Noumedem C, Nzu DDL, et al. Mapping of Schistosomiasis and Soil-Transmitted Helminthiasis in the Regions of Centre, East and West Cameroon. PLoS Negl Trop Dis. 2012;6(3):e1553.

24. Ebai CB, Kimbi HK, Sumbele IUN, Yunga JE, Lehman LG. Efficacy and safety of praziquantel against Schistosoma haematobium in the Ikata-Likoko area of southwest Cameroon. Trop Med Health. 2017;45(1):30.

25. Clinical Overview of Paragonimiasis. Paragonimiasis. CDC, Atlanta. 2024. https://www.cdc.gov/paragonimus/hcp/clinical-overview/index.html? Accessed: 2024 Nov 23.

26. Moyou-Somo R, Kefie-Arrey C, Dreyfuss G, Dumas M. An epidemiological study of pleuropulmonary paragonimiasis among pupils in the peri-urban zone of Kumba town, Meme Division, Cameroon. BMC Public Health. 2003; 3:40.

27. Pourhoseingholi MA, Vahedi M, Rahimzadeh M. Sample size calculation in medical studies. Gastroenterol Hepatol Bed Bench. 2013;6(1):14–7.

28. Kim JS. A Comparison Of Sensitivity On Stool And Sputum Examination For Diagnosis Of Paragonimasis. Kisaengchunghak Chapchi. 1970;8(1):22–4.

29. Nawa Y. Re-emergence of paragonimiasis. Intern Med. 2000; 39:353–4.

30. Narain K, Devi KR, Bhattacharya S, Negmu K, Rajguru SK, Mahanta J. Declining prevalence of pulmonary paragonimiasis following treatment & community education in a remote tribal population of Arunachal Pradesh, India. Indian J Med Res. 2015;141(5):648–52.

31. Eke RA, Nwosu UM, Enwereji EE, Emerole CV. Paragonimiasis Reemergence in Nigeria: Predisposing Factors and Recommendations for Early Intervention and Everlasting Eradication. Int Sch Res Not. 2013;2013(1):257810.

32. Reginald AE, Udochi MN, Ezinne EE, Chima VE. Paragonimiasis Reemergence in Nigeria: Predisposing Factors and Recommendations for Early Intervention and Everlasting Eradication. Hindawi Publ Corp. 2012; 2013:1–4.

33. Ollivier G, Boussinesq M, Albaret JL, Cumberlidge N, Farhati K, Chippaux JP, et al. Etude épidémiologique d’une distomatose à Paragnonimus sp. au Sud-Cameroun. Bull Société Pathol Exot Ses Fil. 1996;88(04):164–9.

34. Kagawa FT. Pulmonary paragonimiasis. Semin Respir Infect. 1997;12(2):149–58.

35. Shin DH, Joo CY. Prevalence of Paragonimus westermani in some Ulchin school children. Acta Paediatr Jpn Overseas Ed. 1990;32(3):269–74.

36. Moyou-Somo R, Tagni-Zukam D. Paragonimiasis in Cameroon: clinicoradiologic features and treatment outcome. Med Trop Rev Corps Sante Colon. 2003;63(2):163–7.

37. Mahajan RC. Paragonimiasis: an emerging public health problem in India. Indian J Med Res. 2005;121(6):716–8.

38. Indrawati I, Chaicumpa W, Setasuban P, Ruangkunaporn Y. Studies on immunodiagnosis of human paragonimiasis and specific antigen of Paragonimus heterotremus. Int J Parasitol. 1991;21(4):395–401.

